# Motor impairment, balance, and muscle coactivation limit the effectiveness of voluntary corrections of asymmetry during walking after stroke

**DOI:** 10.64898/2026.07.27.26359033

**Authors:** Andrian Kuch, Samantha Jeffcoat, Alejandro Aguirre-Ramirez, Hailey Hashiguchi, Evan Shrier, Andrew Hooyman, Nicolas Schweighofer, Carolee Winstein, Alison McKenzie, Natalia Sánchez

## Abstract

**Introduction:** Several gait rehabilitation approaches after stroke rely on explicit feedback to promote task-specific voluntary corrections of walking patterns. While these approaches show effectiveness at a group level, individual responses to voluntary corrections can differ, limiting the benefits and translation of task-specific gait interventions. Our goal is to identify biomechanical, neuromuscular, and cognitive characteristics associated with the ability to perform voluntary corrections of walking using explicit visual feedback in people with chronic stroke.

**Methods:** Twenty-eight individuals with chronic stroke completed a single-session treadmill walking protocol, consisting of baseline walking, a voluntary correction condition guided by real-time visual feedback, and a short retention trial without feedback. Reducing step length asymmetry was used as the target to guide voluntary corrections. Clinical assessments included measures of motor impairment, balance, gait function, cognition, and walking capacity. Muscle coactivation was characterized using dimensionality reduction. Associations of clinical assessments with baseline step length asymmetry and residual error in step length asymmetry during voluntary correction were examined using univariate analyses and multivariate regression with LASSO-based variable selection.

**Results:** Eighteen participants successfully reduced step length asymmetry using visual feedback, while ten participants did not reduce asymmetry. Greater residual asymmetry during voluntary correction was independently associated with greater baseline asymmetry, greater lower extremity motor impairment, reduced balance, and increased paretic muscle coactivation (adjusted R² = 0.46). Neither the direction of asymmetry nor cognitive outcome measures were associated with the ability to correct asymmetry during walking. Immediate retention after feedback removal was limited, with only 4 participants maintaining improvements.

**Discussion:** The ability to perform voluntary corrections of the walking pattern using voluntary corrections after stroke is constrained by motor impairment, balance function, and muscle coactivation. These findings suggest that explicit, feedback-based gait interventions to guide voluntary corrections may benefit individuals with mild to moderate impairments, while individuals with more severe impairments require alternative strategies to guide corrections of the walking pattern.

## 2 Introduction

The ability to perform voluntary corrections of the gait pattern is vital in people post-stroke. On the one hand, voluntary corrections of the walking pattern are essential for safe community ambulation in the presence of obstacles and changes in terrain (1). On the other hand, voluntary corrections of the walking pattern are common in clinical and research interventions. Clinical and research approaches to retrain gait may use feedback to provide individuals with external, explicit cues to perform voluntary corrections (2). Feedback can include verbal instructions (3, 4), real-time biofeedback based on auditory, haptic, or visual cues (2). For example, studies have used visual feedback to target asymmetries in step lengths (5–9), knee and hip joint angles (10), ground reaction forces (11), joint torques (12), or lower limb muscle activation (12). While these approaches have shown group-level improvements in the specific aspects of gait being targeted by voluntary corrections (2), not all individuals are able to perform voluntary corrections of their gait pattern guided by feedback (5, 6).

Post-stroke responses to feedback-based gait rehabilitation may be shaped by three interrelated classes of constraints: biomechanical limitations (13, 14), neural control impairments (15), and cognitive demands (16, 17). Identifying why some individuals can perform voluntary corrections to fulfill task demands while others cannot remains a critical challenge for translating task-specific gait interventions into practice.

Post-stroke walking impairments involve a combination of complex and heterogeneous biomechanical factors (13, 14) resulting in an asymmetric walking pattern (13), which makes asymmetry reduction a common goal of rehabilitation (13, 18). An explanation for the different responses to feedback relates to the targets used to guide voluntary corrections (19) during gait retraining. In the case of using step length asymmetry (SLA) as a target, individuals may either walk with longer paretic or longer non-paretic steps (20). From a biomechanics perspective, asymmetry due to longer paretic steps is associated with paretic kinetics (21), whereas asymmetry due to shorter paretic steps is associated with both paretic and non-paretic kinetics (21), supporting the idea that correcting asymmetry requires different strategies depending on the direction of asymmetry. In fact, our previous work has shown that individuals who walk with longer paretic steps can lengthen non-paretic step lengths to reduce asymmetry, whereas individuals who walk with shorter paretic steps are less able to increase paretic step lengths to reduce asymmetry (9). Although gait biomechanics influence both asymmetry and its correction post-stroke (22), the relationship between clinical characteristics and asymmetry reduction strategies remains unclear and warrants investigation to inform targeted interventions.

Beyond these biomechanical constraints, individual differences in asymmetry reduction may also reflect the neural control mechanisms that enable voluntary corrections guided by feedback. Voluntary corrections rely on cortical neural pathways disrupted by the stroke lesion (23), with studies showing reduced lower extremity isometric torque generation, emergence of joint synergies, and abnormal muscle coactivation in the paretic (24, 25) and non-paretic (26) extremities during voluntary efforts. This abnormal control may also hinder task performance during voluntary correction (26–28), particularly during a dynamic task such as walking, as the voluntary correction needs to be achieved while simultaneously fulfilling the requirements of weight acceptance, single limb support and limb advancement (29). Additionally, balance function which is impaired post-stroke (30) may also limit the ability to perform voluntary corrections (31, 32). Thus, lower-limb weakness (25, 33), abnormal limb synergies (34), and impaired postural control (33, 35) can constrain the degrees of freedom available for voluntary modification of gait.

Cognitive factors may also influence voluntary corrections of gait post-stroke. Cognitive impairment impairs executive function, visuospatial skills, memory, or attention (36), which affects motor control and rehabilitation (16, 17). These cognitive deficits may therefore compromise the ability to process and act on external cues, as explicit feedback requires attention, interpretation of sensory information, and integration of task goals into motor planning (37). However, it remains unclear whether global measures of cognitive impairment are associated with the ability to perform voluntary gait corrections using visual feedback, or whether motor and functional impairment have a more dominant role.

In this study, we investigated the biomechanical, neuromuscular, and cognitive factors associated with the ability to perform voluntary corrections during gait in people with chronic stroke. We used voluntary correction of step length asymmetry guided by visual feedback. We collected clinical assessments, including measures of sensorimotor impairment, balance and postural control, and cognition, as well as measures of walking function and neuromuscular control during walking. We hypothesize that an individual’s ability to reduce SLA is associated with a combination of gait function, balance function, direction of step length asymmetry, lower-limb motor impairment, and cognitive function. Our results will provide insights into the clinical characteristics of individuals who might benefit from explicit feedback to guide voluntary corrections in clinic and research studies, to inform the personalization of task-specific gait interventions, and support more effective translation of rehabilitation strategies to community ambulation after stroke.

## 3 Materials and Methods

We recruited 28 individuals with chronic stroke (Table 1). Inclusion criteria were: (1) a single documented cerebrovascular accident, (2) self-reported unilateral paresis, (3) no other neuromuscular conditions affecting walking, (4) ability to walk 5 minutes on a treadmill without handrails; use of an ankle-foot orthosis or brace was permitted, (5) no hemispatial neglect as assessed using the clock drawing test (38), (6) ability to provide informed consent. The Chapman University Institutional Review Board number IRB-23-57 approved the study, and all participants provided written informed consent before testing.

**Table 1:**
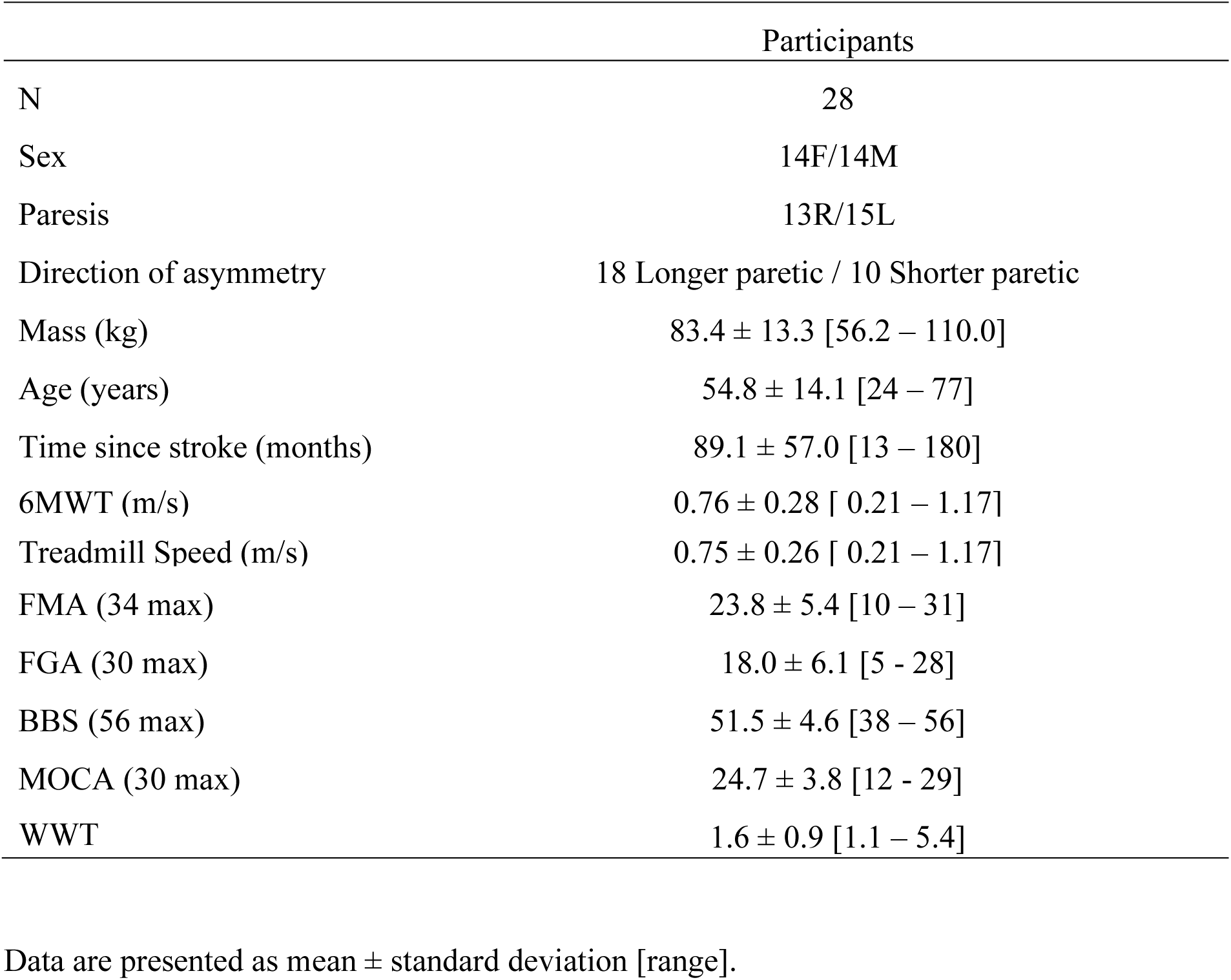
Demographics.

### 3.1 Experimental protocol

Data were collected in the Gait Behavior Lab at Chapman University between August 2023 and May 2025. We collected the following clinical scores: 6-minute walk test (6MWT) (39), lower extremity motor portion of the Fugl-Meyer assessment (FMA) (40), Berg Balance Scale (BBS) (41), Functional Gait Assessment (42), Montreal Cognitive Assessment (43), and Walking While Talking test (WWT) (44). In the WWT test, participants walked at a comfortable speed, and then while reciting alternate letters of the alphabet. We expressed WWT time as 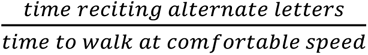.

We collected experimental data in a Gait Real-time Analysis Interactive Lab (GRAIL), equipped with an 180-degree screen to provide visual feedback. The lab is equipped with 10 motion capture cameras, sampling at 100 Hz, and an instrumented treadmill, sampling at 1000 Hz. We placed 26 reflective markers on bony landmarks using a modified Human Body Model 2 (45) for the lower limbs and trunk (Figure 1). We collected muscle activations using surface electromyography (EMG) sampling at 2000 Hz for the Vastus Medialis, Rectus Femoris, Biceps Femoris long head, Semitendinosus, Tibialis Anterior, Soleus, and Lateral Gastrocnemius, bilaterally (Figure 1).

**Figure 1:**
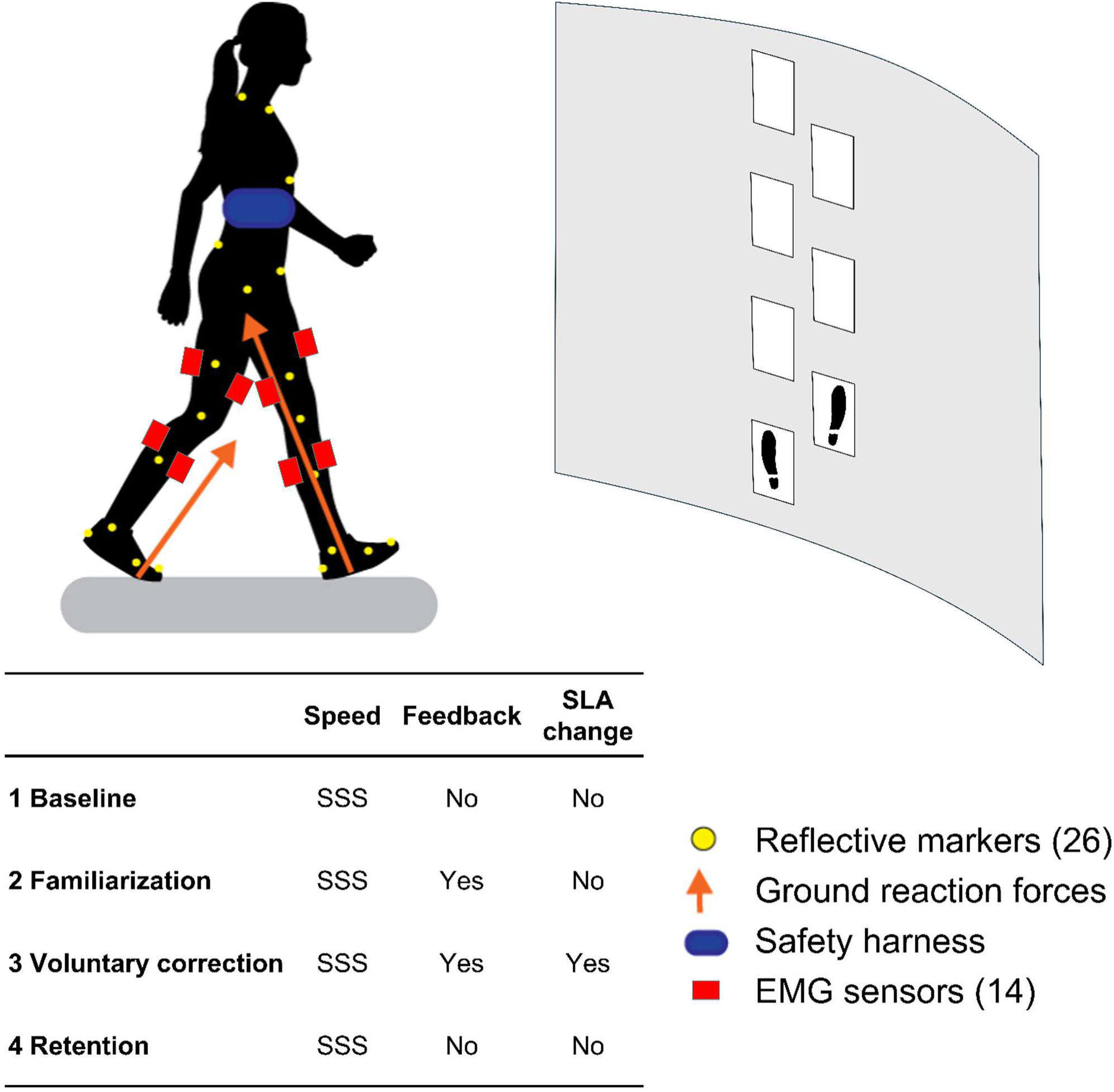
Study protocol. 1) Participants first walked at their self-selected speed (SSS) for a baseline walking trial. 2) They familiarized themselves with the visual feedback of steps targets, without modifying their step length asymmetry (SLA). 3) Voluntary correction trial with visual feedback of step targets at self-selected speed to promote symmetric step lengths. 4) Retention trial without visual feedback

For all trials, we set the treadmill speed at the participant’s self-selected speed (SSS), or at least 85% of the 6MWT overground speed if they were unable to maintain the SSS. Participants wore an overhead safety harness to prevent falls, without providing any body weight support (Figure 1).

Data collection began with a 2-minute baseline trial at SSS to collect participants’ baseline step length asymmetry (|*SLA*_SSS_|). Participants then familiarized themselves with the visual feedback for 2 minutes: footprints on a screen, updated in real time based on heel marker positions, and tiles indicating the step length target locations for the following steps (Figure 1). During familiarization, the target step lengths matched baseline values without modifying SLA.

For the voluntary correction trial, we provided visual feedback of step targets to guide symmetric step lengths for 2 minutes and measured the magnitude of remaining asymmetry during voluntary correction (|*SLA*_SFM–FBK_|). Step-length targets were set to match the longer step measured during baseline while maintaining step width. While this approach impacts cadence, it prevents the unwanted effect of shortening step lengths to achieve symmetry(21). Immediately after, participants walked 2 minutes without visual feedback in a retention trial as we measured retention SLA (|*SLA*_RET_|).

Step length was calculated as the anteroposterior difference between heel markers at each respective limb heel strike, in mm. SLA was defined as the absolute difference between the paretic and non-paretic step lengths to account for individuals taking longer paretic or non-paretic steps. The direction of SLA was coded as a binary variable: 1 for shorter paretic steps, and 2 for longer paretic steps. SLA averages and SLA direction were obtained for the last 20 strides of the baseline and visual feedback trials, and the first 20 strides of the retention trial.

### 3.2 Data processing

We processed all data using custom MATLAB 2023b code. We filtered kinematic data using a 4^th^-order Butterworth lowpass filter with a 10Hz cutoff, and kinetic data using a 4th-order Butterworth lowpass filter with a 20Hz cutoff. We combined kinematic and kinetic data to detect heel strikes and segment walking trials into strides. EMG processing was done as in Sanchez 2025(46). We quantified muscle coactivation using the Dynamic Motor Control index (WalkDMC). WalkDMC(47) that quantifies overall muscle coactivation relative (48–50), validated in people post-stroke to characterize paretic and non-paretic (46). A WalkDMC of 100 represents the coactivation of neurotypical controls; a value below 100 indicates an increase in coactivation; a value above 100 indicates a decrease in coactivation.

### 3.3 Statistics

#### 3.3.1 Multivariate association between residual asymmetry during voluntary corrections and clinical characteristics association between asymmetry and clinical characteristics

We used a two-stage modeling strategy to identify clinical characteristics associated with asymmetry during voluntary corrections |*SLA*_SFM–FBK_|. First, we performed Lasso regression (51) to model |*SLA*_SFM–FBK_| with Leave-One-Out Cross Validation to prevent model overfitting (52). To improve stability, we repeated the Lasso 1,000 times using different random seeds and used the average regularization parameter *λ* across all iterations (52). Second, since coefficients from Lasso represent penalized estimates, we re-fitted a linear regression model with the identified subset of variables to obtain standardized regression coefficients. We evaluated linearity and homoscedasticity using residuals-versus-fitted plots, normality of residuals with Q–Q plots, and multicollinearity using variance inflation factors (VIF).

#### 3.3.2 Multivariate association between baseline asymmetry and clinical characteristics

To identify clinical characteristics associated with baseline step length asymmetry, we followed the same procedure as described above, but using baseline step length asymmetry as a response variable.

#### 3.3.3 Univariate association between asymmetry and clinical characteristics

We assessed the association between baseline asymmetry and participants’ clinical (age, time since stroke, measures of motor and cognitive impairment), and baseline walking characteristics (SSS, direction of SLA, bilateral muscle coactivation) to better understand how clinical factors relate to asymmetry. We assessed bivariate data normality using Mardia’s multivariate normality test (53). We computed Pearson’s correlation coefficient for all associations, except for the association between baseline asymmetry and the direction of asymmetry, where we computed Spearman’s rank correlation coefficient. We used a False Discovery Rate Benjamini-Hochberg procedure to adjust p-values for multiple comparisons (54). We present the unadjusted and adjusted significance for all correlations.

#### 3.3.4 Effect of visual feedback on asymmetry and coactivation

To assess the effect of walking condition on step length asymmetry, we fitted a linear mixed-effects model, with SLA as the dependent variable and walking condition (baseline, voluntary correction, retention) as a fixed effect, and a random 1|ID intercept.

To assess the effect of walking condition on coactivation in the paretic and non-paretic side, we fitted linear mixed-effects models, with either paretic or non-paretic WalkDMC as the dependent variable and walking condition as a fixed effect, and a random 1|ID intercept.

Model assumptions were assessed using residual-versus-fitted plots to assess linearity and homoscedasticity, and Q–Q plots to evaluate residual normality. The normality of random intercepts was examined with Q–Q plots, and variance components were inspected. Post-hoc comparisons between walking conditions were conducted using estimated marginal means with False Discovery Rate Benjamini-Hochberg corrections. The significance level for all tests was set at α = 0.05. Statistical analyses were performed in R (version 4.3.2). Descriptive statistics are presented as mean ± SD, and regression coefficients as mean (SE).

#### 3.3.5 Sample size

The sample size of 28 participants was chosen to provide adequate power to detect correlations of about |r| ≥ 0.5 at a significance level α = 0.05 under a bivariate normal model, which were considered the smallest effects to be clinically meaningful (55).

## 4 Results

### 4.1 Residual asymmetry during voluntary correction was associated with baseline asymmetry, motor impairment, balance, and paretic coactivation

The residual asymmetry during voluntary correction |*SLA*_SFM–FBK_| LASSO model retained four variables: |*SLA*_SSS_|, FMA, BBS, and paretic WalkDMC (Table 2). Regression assumptions were met, and all VIFs < 2. From the unstandardized model (Table 3, model 1), each 1-point increase in FMA, BBS, and paretic WalkDMC was associated with a 2.0 mm, 1.0 mm, and 0.49 mm reduction in |*SLA*_SFM–FBK_| respectively, while a 1 mm increase in |*SLA*_SSS_| was associated with a 0.44 mm increase of |*SLA*_SFM–FBK_|. From the standardized model (Table 2, model 1, Figure 2), |*SLA*_SSS_| was the most influential variable (β=0.51, SE=0.18, 95%CI= [0.16, 0.86]), followed by FMA (β=-0.20, SE=0.18, 95%CI= [−0.56, 0.16]), BBS (β=−0.09, SE=0.17, 95%CI= [−0.42, 0.25]), and paretic WalkDMC (β=−0.08, SE=0.19, 95%CI= [−0.45, 0.29]). Since predictors were selected using LASSO, classical p-values were not interpreted to avoid post-selection inference bias (56).

**Table 2:** Model to predict asymmetry during voluntary correction Regression Coefficients.

**Regression Coefficients**
| *Dependent variable:* |  |  |
| --- | --- | --- |
| <u>$\vert SLA_{SYM-FBK}\vert $</u> |  |  |
|  | Unstandardized | Standardized |
| FMA | -2.00 (1.81) | -0.20 (0.18) |
| BBS | -1.01 (1.99) | -0.09 (0.17) |
| $\vert SLA_{SSS}\vert $ | 0.44 (0.15) | 0.51 (0.18) |
| $P\ WalkDMC_{SYM-FBK}$ | -0.49 (1.18) | -0.08 (0.19) |
| Constant | 161.12 (121.99) |  |
| Observations | 28 |  |
| R<sup>2</sup> | 0.54 |  |
| Adjusted R<sup>2</sup> | 0.46 |  |
| Residual Std. Error (df = 25) | 39.59 |  |
| F Statistic (df = 2; 25) | 6.67 |  |

Estimated coefficients are presented as mean (standard error). Since predictors were selected using
LASSO, classical p-values were not interpreted to avoid post-selection inference bias

**Figure 2:**
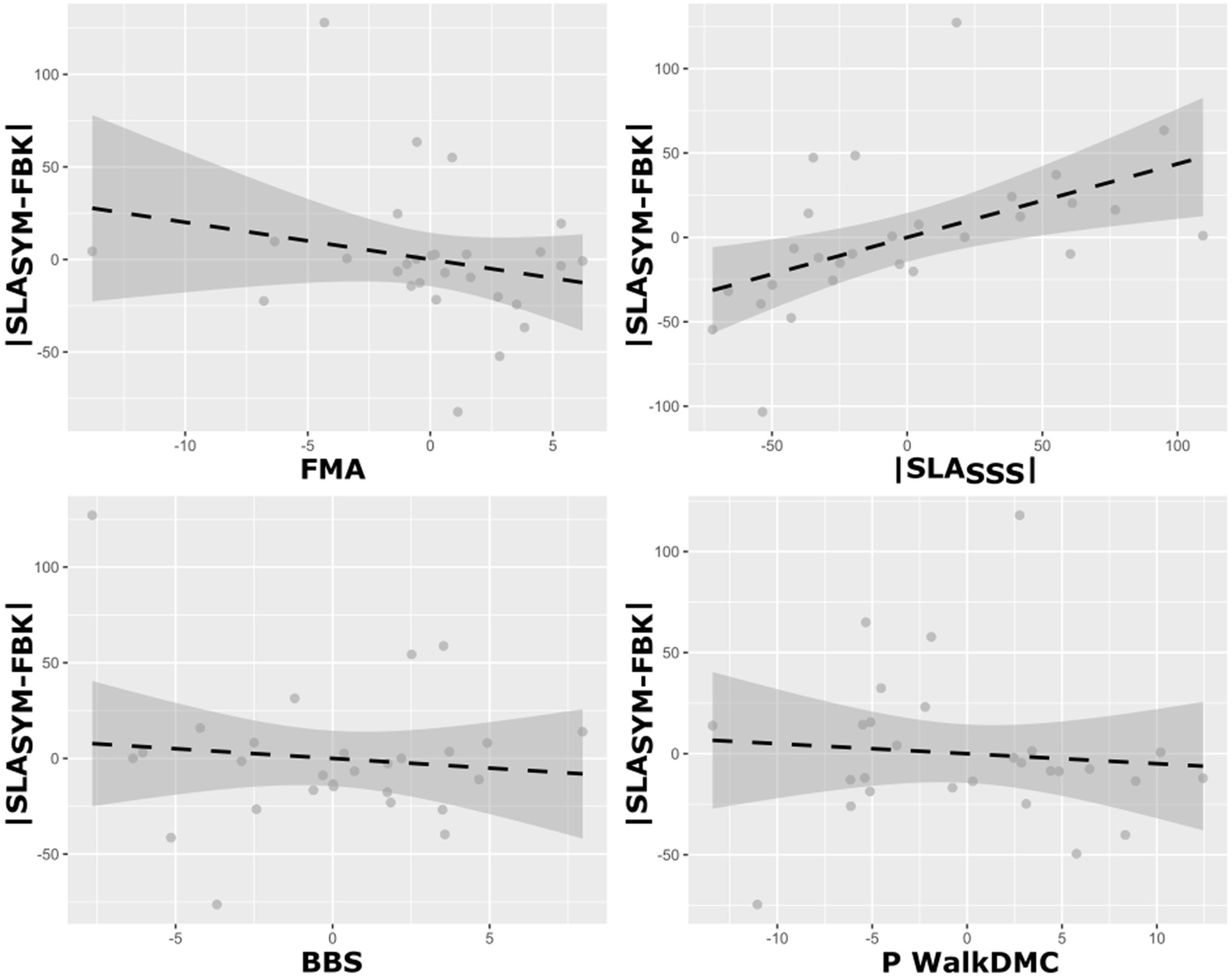
Partial regression plots illustrating the unique contribution of Fugl-Meyer (FMA), Berg Balance Scale (BSS), baseline step length asymmetry |*SLA*_SSS_|, and paretic coactivation (P WalkDMC) to step length asymmetry during voluntary correction |*SLA*_SFM–F_ |, while maintaining the other predictors constant.

**Table 3:**
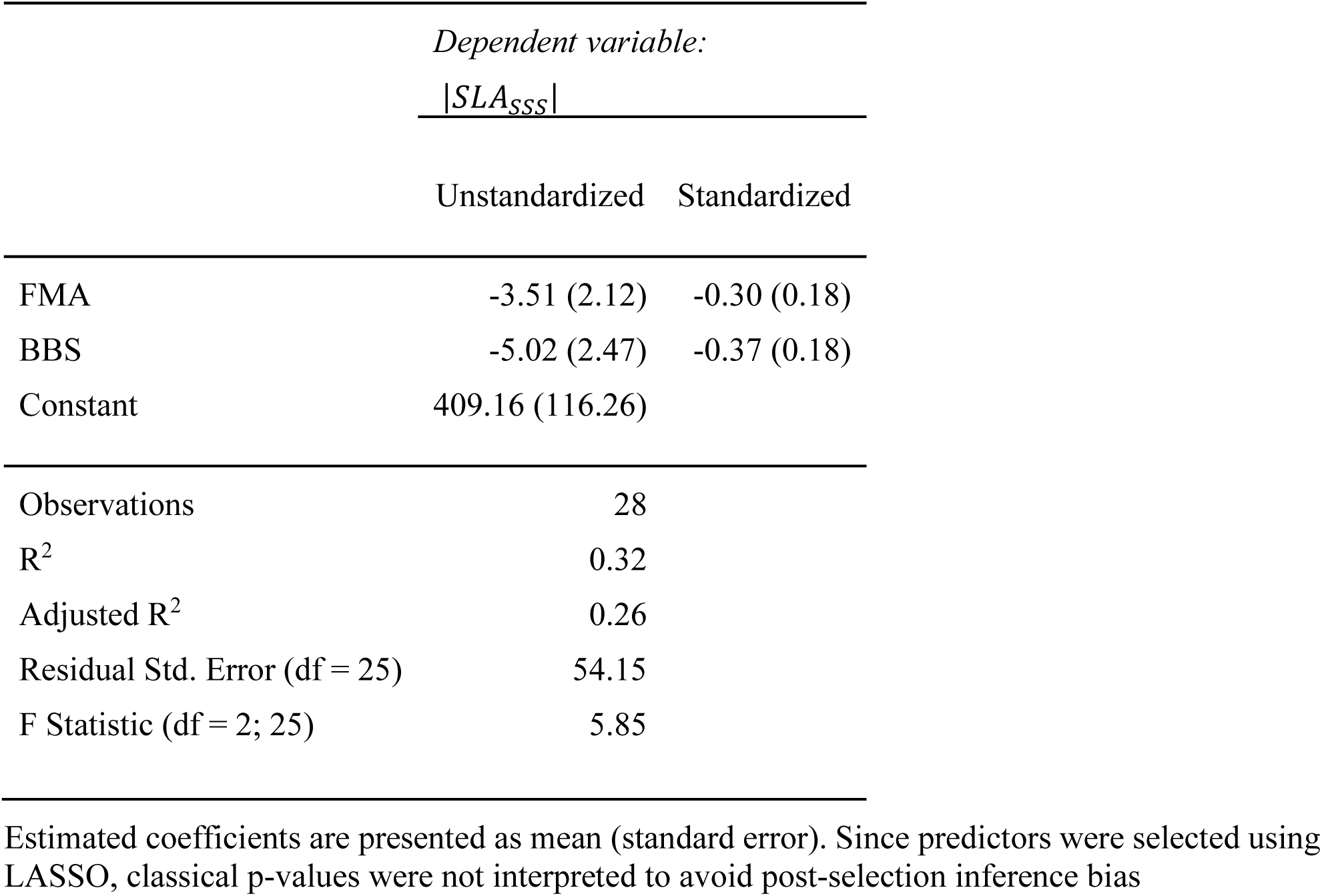
Model to predict baseline asymmetry.

### 4.2 Baseline asymmetry was associated with balance and motor impairment

The baseline asymmetry |*SLA*_SSS_| LASSO model retained two variables: BBS and FMA (Table 3). Regression assumptions were met, and all VIF < 2. From the unstandardized model (Table 3, model 1), each 1-point increase in FMA and BBS was associated with a 3.5 mm and 5.0 mm reduction in |*SLA*_SSS_|, respectively. From the standardized model (Table 3, model 2, Figure 3), BBS (β=−0.37, SE=0.18, 95%CI= [−0.73, −0.01]) showed a slightly stronger association with |*SLA*_SSS_| than FMA (β=−0.30, SE=0.18, 95%CI= [−0.66, 0.06]).

**Figure 3:**
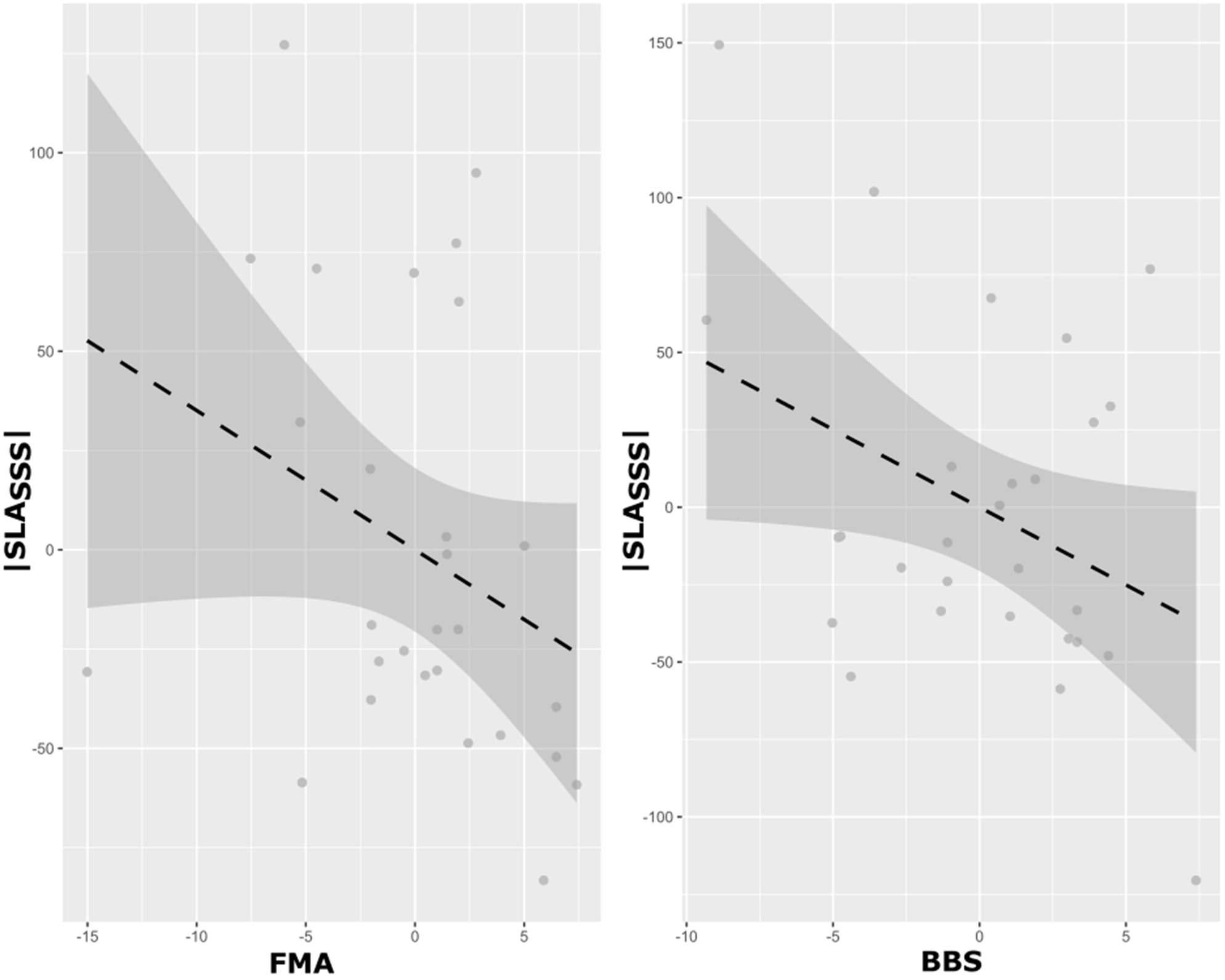
Partial regression plots illustrating the unique contribution of Fugl-Meyer (FMA), Berg Balance Scale (BSS) to baseline step length asymmetry |*SLA*_SSS_|, while maintaining the other predictor constant.

### 4.3 Univariate association between baseline asymmetry and clinical characteristics

At a group level, we observed a baseline step length asymmetry of |*SLA*_SSS_| = 67.0 ± 63.1 mm. We observed that 10/28 participants had shorter paretic steps and 18/28 had longer paretic steps. We observed moderate correlations between |*SLA*_SSS_| and clinical characteristics (Figure 4): FMA (r = −0.45, *p* = 0.015, *p*_adj_ = 0.06), BBS (r = −0.49, *p* = 0.01, *p*_adj_ = 0.06), SSS (r = −0.45, *p* = 0.02, *p*_adj_ = 0.06), FGA (r = −0.39, *p* = 0.04, *p*_adj_ = 0.09), and paretic WalkDMC (r = −0.40, *p* = 0.04, *p*_adj_ = 0.09). We observed no significant correlations for time since stroke (r = −0.23, *p* = 0.24, *p*_adj_ = 0.37) and WWT (r = −0.33, *p* = 0.09, *p*_adj_ = 0.16).

**Figure 4:**
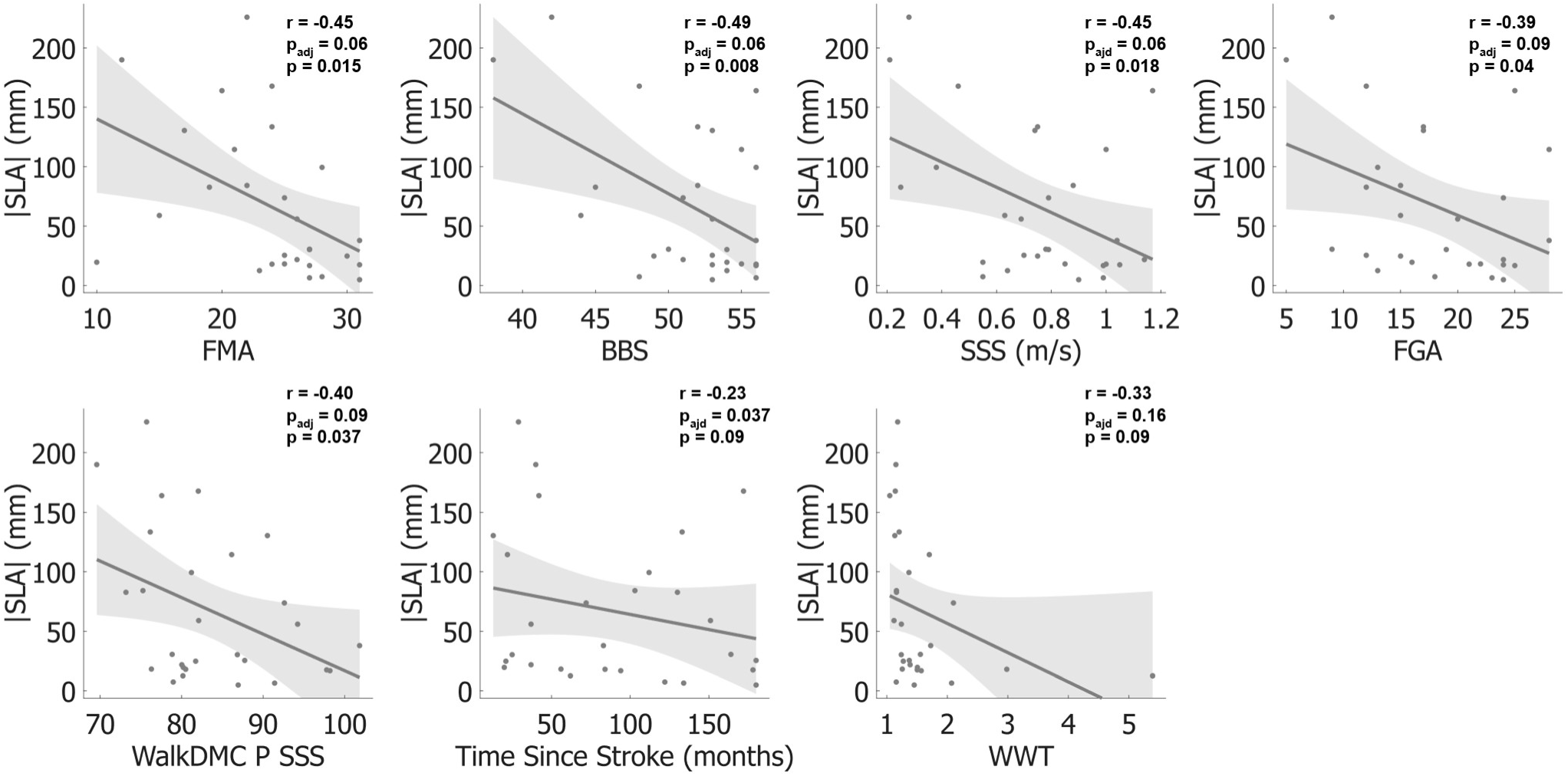
Correlations of baseline step length asymmetry with Fugl-Meyer (FMA), Berg Balance Scale (BBS), self-selected speed (SSS), Functional Gait Assessment (FGA), paretic (WalkDMC P), Time since stroke, and Walking While Talking (WWT). Unadjusted and FDR adjusted p-values are presented.

Correlations between |*SLA*_SSS_| and age (r = 0.02, *p* = 0.91, *p*_adj_ = 0.91), MOCA (r = 0.12, *p* = 0.54, *p*_adj_ = 0.59), non-paretic WalkDMC (r = −0.20, *p* = 0.31, *p*_adj_ = 0.42) and direction of asymmetry (Spearman *ρ* = 0.45, *p* = 0.37, *p*_adj_ = 0.45) were not significant with p>0.250.

### 4.4 Individuals reduce asymmetry with feedback, but do not retain asymmetry reductions

We observed that 18 of 28 individuals reduced SLA with lower asymmetry during voluntary correction than during baseline walking (|*SLA*_SFM–FBK_| < |*SLA*_SSS_|, Figure 5), including 11 with longer paretic and 7 with shorter paretic steps at baseline. Of the 10 participants who did not reduce asymmetry, 9 had a baseline SLA of 14.5 ± 7.3 mm (n=9), which falls within the value of neurotypical controls (57). An outlier participant with a baseline SLA of 190.0 mm, increased their asymmetry with feedback to 265 mm, and to 313 mm during retention. After removing this outlier, |*SLA*_SSS_| was significantly lower (*p* = 0.005) in the 9 non-responders compared to the 18 responders (83.8 ± 61.8 mm), suggesting a ceiling effect in the non-responders. Among the 18 responders, only 4 retained the reduced SLA within 15 mm once the feedback was removed (Figure 5).

**Figure 5:**
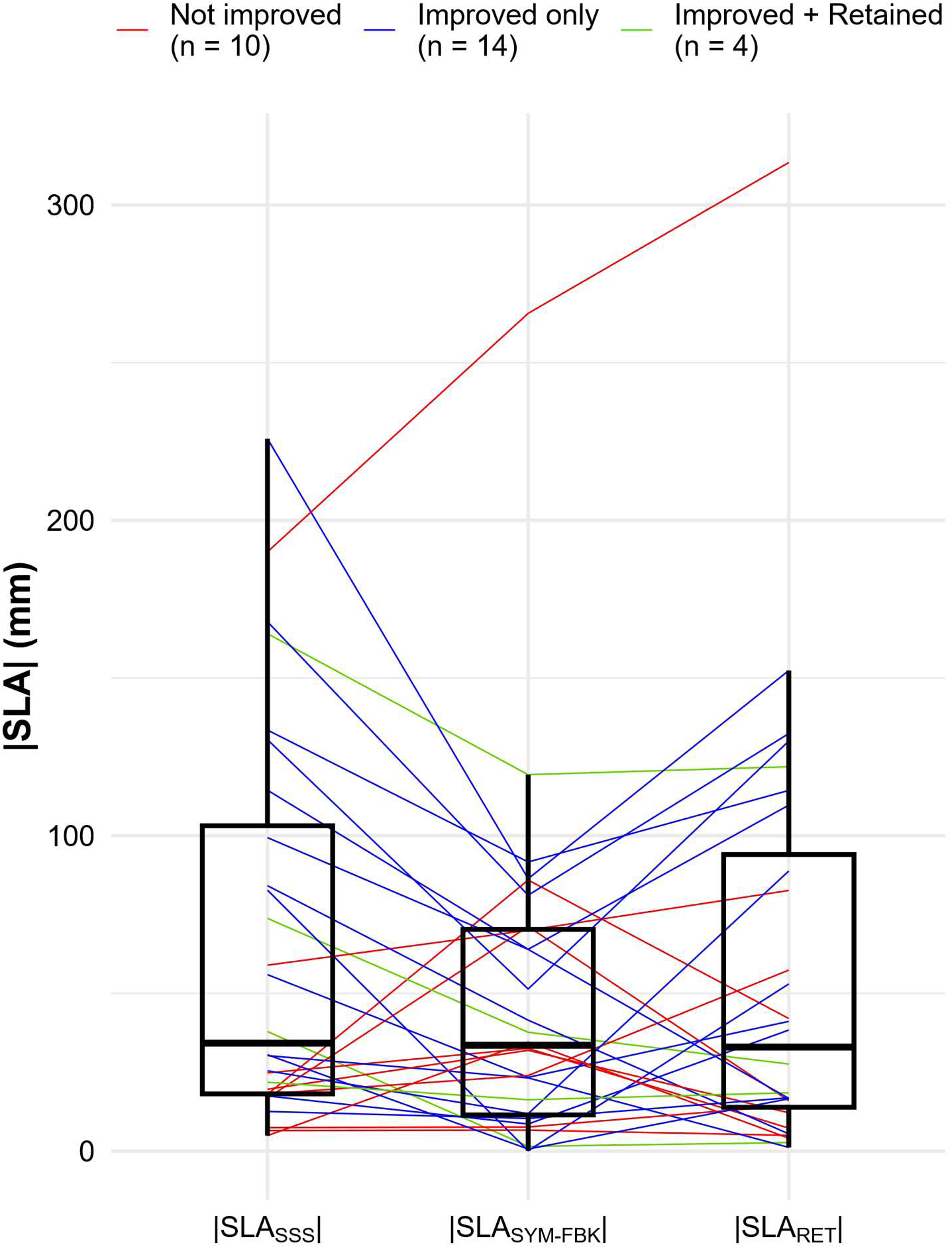
Magnitude of step length asymmetry across the three walking conditions (baseline |SLA_SSS |, voluntary correction |SLA_(SYM-FBK) |, retention |SLA_RET |). Red line: individuals who did not reduce step length asymmetry during voluntary correction trial and thus did not improve. Blue line: individuals who reduced step length asymmetry during the voluntary correction trial but did not retain the step length asymmetry immediately after. Green line: individuals who reduced step length asymmetry during the voluntary correction trial and retained the step length asymmetry immediately after.

For the full sample, the linear mixed-effects model showed no effect of walking condition on SLA (F(2,54)=2.59, p=0.08, Figure 5). After excluding the outlier, we observed a significant main effect of walking condition on SLA (F(2,52) = 4.3, p =0.02). Post-hoc comparisons using Bonferroni-adjusted estimated marginal means revealed that |*SLA*_SSS_| was significantly higher (62.3 ± 9.3 mm) than |*SLA*_FBK–SFM_| (40.6 ± 9.3 mm; p = 0.016). |*SLA*_RET_| (49.2 ± 9.3 mm) did not differ significantly from |*SLA*_SSS_| (p = 0.25) or from |*SLA*_FBK–SFM_| (p = 0.77).

Both linear mixed-effects models to assess differences in paretic coactivation (*F*(2, 54) = 0.75, *p* = 0.48) and non-paretic coactivation (*F*(2, 54) = 2.07, *p* = 0.14) across walking conditions were not significant. Paretic WalkDMC was 83.7 ± 1.7 during baseline, 84.1 ± 1.7 during voluntary correction, and 84.9 ± 1.7 during retention (Figure 6). Non-paretic WalkDMC was 88.3 ± 1.6 during baseline, 86.9 ± 1.6 during voluntary correction, and 88.5 ± 1.6 during retention (Figure 6).

**Figure 6:**
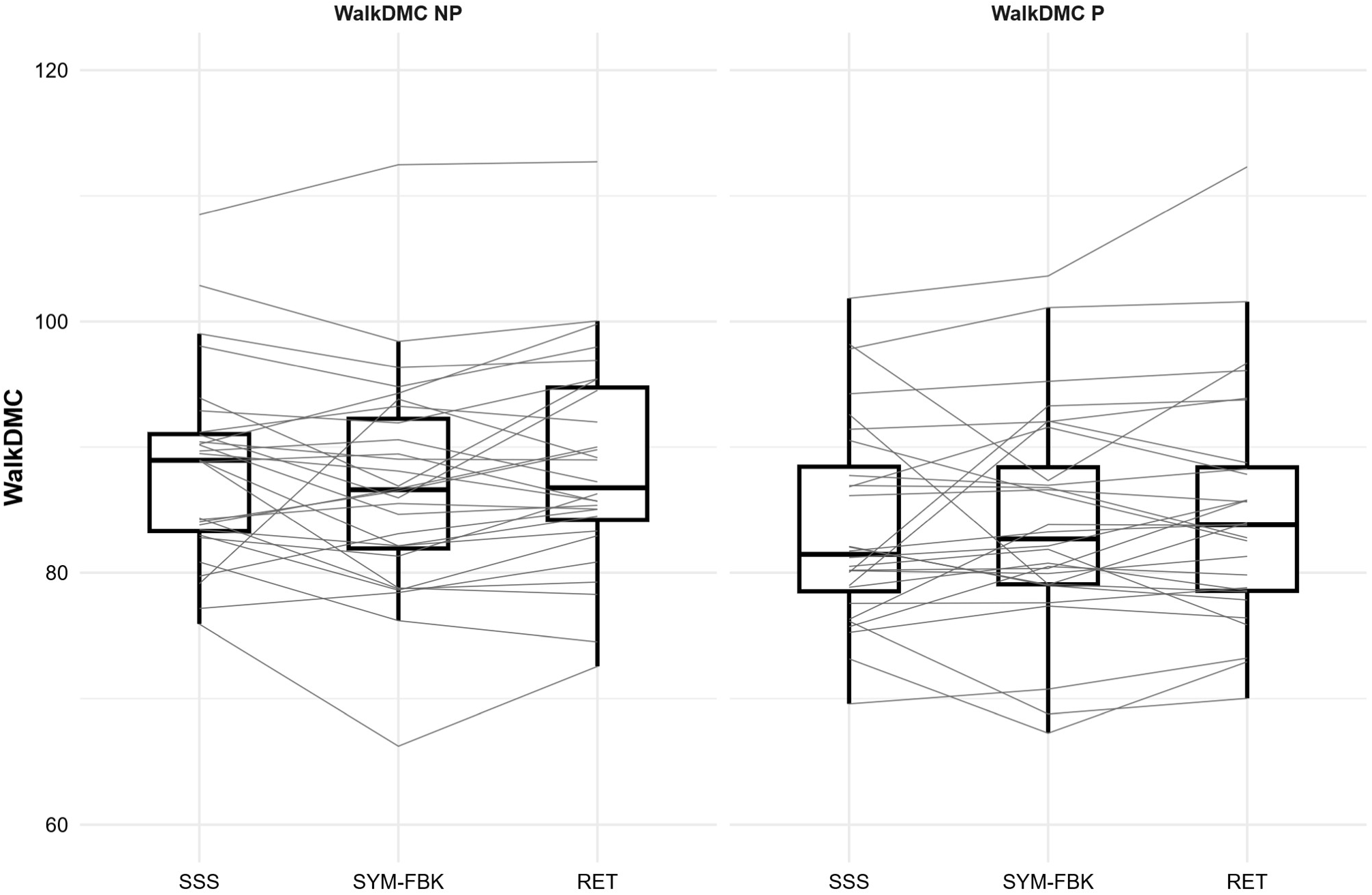
Muscle coactivation WalkDMC across the three walking conditions (baseline SSS, voluntary correction SYM-FBK, retention RET) in the non-paretic NP side (left) and paretic P side (right).

## 5 Discussion

Task-specific clinical and research gait retraining strategies commonly use feedback to cue voluntary correction of the walking pattern post-stroke (2), yet the ability to modify specific aspects of gait varies across individuals (5, 6, 9). In this study, we used voluntary correction of step length asymmetry to examine the clinical and neuromuscular constraints that limit this capacity. Using visual feedback to guide symmetrical step lengths, we showed that individuals with greater motor and balance impairments, as measured by the FMA and the BBS, were less able to reduce asymmetry. Individuals who showed higher paretic coactivation (lower WalkDMC) were also less able to reduce asymmetry. Contrary to our hypothesis, neither direction of asymmetry nor cognitive impairment was associated with the ability to reduce asymmetry guided by visual feedback. These results provide important insights into why explicit, feedback-based gait interventions to guide voluntary corrections of the walking pattern show heterogeneous responses across individuals and have direct implications to inform which individuals may benefit from explicit gait interventions.

### 5.1 Voluntary gait corrections reflect motor capacity

Our results show that baseline SLA was the strongest clinical characteristic associated with residual asymmetry during voluntary correction. Its standardized regression coefficient was 0.51, compared to 0.2 for FMA, 0.09 for BBS, and 0.08 for paretic WalkDMC. One explanation is that participants with more asymmetry have more room for improvement, while those with lower asymmetry have little left to reduce. However, one outlier seems to contradict this: with a baseline SLA of 190 mm, this participant showed no improvement in asymmetry. Notably, this participant had severe impairment (SSS=0.21 m/s, FMA=12, BSS=38, and FGA=4). Thus, even if asymmetric, participants who are more severely impaired may be unable to perform voluntary corrections during walking and might require more implicit types of feedback to retrain walking patterns.

Our findings also show that motor impairment and balance function independently limited the ability to reduce SLA, even after accounting for baseline SLA. Participants with lower Fugl-Meyer and Berg Balance Scale scores exhibited greater residual SLA during feedback. These findings align with prior work showing that impaired balance (8, 58) and motor control (22) are associated with asymmetry after stroke. Importantly, our results extend this knowledge by demonstrating that these impairments also limit the ability to voluntarily modify asymmetry. This has significant implications for task-specific training paradigms which require more intact motor impairment and balance to allow task practice that would promote locomotor learning.

Our findings also show that coactivation limited the ability to reduce SLA. Contrary to previous studies suggesting that increased voluntary effort during lower extremity tasks post-stroke is achieved through heightened muscle coactivation (25, 26), we found no significant differences in coactivation between baseline walking and voluntary corrections. This does not imply that coactivation does not influence voluntary corrections during walking. Here, paretic coactivation limited the reductions in asymmetry using visual feedback, which might indicate that abnormal neuromuscular control and muscle synergies may limit task performance during a voluntary gait correction task (50, 59, 60).

Surprisingly, despite FGA being a predictor of community ambulation(61) and a measure of dynamic balance during functional walking tasks (42), we did not identify FGA as a predictor of the ability to perform voluntary corrections during gait. In individuals post-stroke, FGA is strongly associated with walking speed and BBS (8, 62). Given this overlap, the absence of FGA in the final models might be explained by more by collinearity and less by its construct validity: FGA’s shared variance with BBS, walking speed and other retained predictors potentially led LASSO to drop it, because it did not contribute additional unique explanatory power beyond the variables already included (52).

### 5.2 Voluntary gait corrections were not related to cognitive performance

We observed that cognitive assessments (MOCA, WWT) were not associated with SLA during voluntary correction. Cognitive impairment post-stroke may disrupt executive function, visuospatial skills, memory, or attention (36), and such deficits may in turn hinder rehabilitation (16, 17). More specifically, individuals with better visuospatial skills and locomotor function perform better in a visual feedback gait retraining task (63). An explanation could be that MOCA is a general screening tool, while WWT primarily screens the attention domain(44), making both not specific enough to assess visuospatial skills (17). General cognitive assessments might also miss dynamic and task-related changes (64). Here, our results suggest that cognitive impairment did not limit the ability to modify the gait pattern using a simple feedback task, and for this task, performance was more related to motor than cognitive impairments.

### 5.3 Baseline asymmetry is related to impairment and balance

Our results suggest that baseline SLA is characterized by an individual’s functional abilities and post-stroke locomotor function. Participants with better balance and less motor impairment walked more symmetrically, consistent with previous research showing that poor balance (8, 58), or impairment severity (22) were related to asymmetries post-stroke. Walking speed was not directly associated with gait asymmetry, since a similar gait speed might be achieved using distinct strategies (22, 57, 65). Overall, our results indicate that the combined balance and motor impairments shape baseline asymmetry.

### 5.4 Limited retention suggests no learning

Although most responders reduced task error during visual feedback, only a small subset retained improvements once feedback was removed. This limited retention suggests that explicit feedback-driven corrections for a short period of two minutes may not strongly engage motor learning.

### 5.5 Study limitations

We included ambulatory participants, so our results might not apply to non-functional ambulators or those with more severe impairment, as shown by our outlier. Testing a single visual feedback type limits generalization to other visual, auditory, or haptic feedback. Our sample size provided sufficient power for moderate effects, so smaller but meaningful associations may have gone undetected in complex regression models. We assessed only immediate responses, and retention occurred in just four participants, suggesting that the training dose was insufficient, the timing of the retention did not allow enough time for consolidation, or that the feedback did not engage motor learning mechanisms.

## 6 Conclusion

Voluntary modifications of walking are essential for effective gait rehabilitation and safe community ambulation after stroke. Our findings demonstrate that the ability to perform voluntary gait corrections using explicit visual feedback is constrained by motor impairment severity, balance impairment, and abnormal neuromuscular control. These findings highlight the need to develop other types of gait retraining approaches, such as implicit learning or the use of alternative types of feedback to promote walking modifications in individuals with more severe impairment. Our results indicate that explicit, task-specific gait interventions should therefore not be applied uniformly across individuals post-stroke as they are most beneficial for individuals with mild to moderate impairment.

## 8 Author Contributions

AK: Conceptualization, Investigation, Formal analysis, Software, Visualization, Methodology, Writing – original draft, SJ: Data curation, Investigation, Project administration, Writing – review & editing, AAR: Data curation, Investigation, Writing – original draft, HH: Data curation, Investigation, Writing – original draft, ES: Data curation, Investigation, Writing – original draft, AH: Conceptualization, Methodology, Writing – review & editing, NSc: Conceptualization, Methodology, Writing – review & editing, CW: Conceptualization, Methodology, Writing – review & editing, AM: Writing – review & editing, NSa: Conceptualization, Funding acquisition, Methodology, Supervision, Writing – review & editing.

## 10 Acknowledgments

The authors would like to thank Dr. Ryan Roemmich for insightful and constructive comments on the manuscript. The authors would like to thank all participants in our study.

## 12 Data availability statement

The datasets analyzed for this study can be found in an Open Science Framework repository at: https://osf.io/fc284/overview.

## References

1. Hawkins KA, Clark DJ, Balasubramanian CK, Fox EJ. Walking on uneven terrain in healthy adults and the implications for people after stroke. NeuroRehabilitation. 2017;41(4):765–74.

2. Spencer J, Wolf SL, Kesar TM. Biofeedback for Post-stroke Gait Retraining: A Review of Current Evidence and Future Research Directions in the Context of Emerging Technologies. Frontiers in Neurology: Frontiers Media S.A.; 2021.

3. Parker CJ, Guerin H, Buchanan B, Lewek MD. Targeted verbal cues can immediately alter gait following stroke. Topics in Stroke Rehabilitation. 2022;29(5):382–91.

4. Tate JJ, Milner CE. Real-Time Kinematic, Temporospatial, and Kinetic Biofeedback During Gait Retraining in Patients: A Systematic Review. 2010.

5. Drużbicki M, Guzik A, Przysada G, Kwolek A, Brzozowska-Magoń A, Sobolewski M. Changes in gait symmetry after training on a treadmill with biofeedback in chronic stroke patients: A 6-month follow-up from a randomized controlled trial. Medical Science Monitor. 2016;22:4859–68.

6. Druzbicki M, Przysada G, Guzik A, Brzozowska-Magoń A, Kołodziej K, Wolan-Nieroda A, et al. The efficacy of gait training using a body weight support treadmill and visual biofeedback in patients with subacute stroke: A randomized controlled trial. BioMed Research International. 2018;2018.

7. Padmanabhan P, Rao KS, Gulhar S, Cherry-Allen KM, Leech KA, Roemmich RT. Persons post-stroke improve step length symmetry by walking asymmetrically. Journal of NeuroEngineering and Rehabilitation. 2020;17(1).

8. Park S, Liu C, Sánchez N, Tilson JK, Mulroy SJ, Finley JM. Using Biofeedback to Reduce Step Length Asymmetry Impairs Dynamic Balance in People Poststroke. Neurorehabilitation and Neural Repair. 2021;35(8):738–49.

9. Sánchez N, Finley JM. Individual Differences in Locomotor Function Predict the Capacity to Reduce Asymmetry and Modify the Energetic Cost of Walking Poststroke. Neurorehabilitation and Neural Repair. 2018;32(8):701–13.

10. Day KA, Cherry-Allen KM, Bastian AJ. Individualized feedback to change multiple gait deficits in chronic stroke. Journal of NeuroEngineering and Rehabilitation. 2019;16(1).

11. Genthe K, Schenck C, Eicholtz S, Zajac-Cox L, Wolf S, Kesar TM. Effects of real-time gait biofeedback on paretic propulsion and gait biomechanics in individuals post-stroke. Topics in Stroke Rehabilitation. 2018;25(3):186–93.

12. Tamburella F, Moreno JC, Herrera Valenzuela DS, Pisotta I, Iosa M, Cincotti F, et al. Influences of the biofeedback content on robotic post-stroke gait rehabilitation: Electromyographic vs joint torque biofeedback. Journal of NeuroEngineering and Rehabilitation. 2019;16(1).

13. Olney SJ, Richards C. Hemiparetic gait following stroke. Part I: Characteristics. Gait & Posture. 1996;4(2):136–48.

14. Hendrickson J, Patterson KK, Inness EL, McIlroy WE, Mansfield A. Relationship between asymmetry of quiet standing balance control and walking post-stroke. Gait and Posture. 2014;39(1):177–81.

15. Malone LA, Bastian AJ. Thinking about walking: Effects of conscious correction versus distraction on locomotor adaptation. Journal of Neurophysiology. 2010;103(4):1954–62.

16. Lingo VanGilder J, Hooyman A, Peterson DS, Schaefer SY. Post-Stroke Cognitive Impairments and Responsiveness to Motor Rehabilitation: A Review. Current Physical Medicine and Rehabilitation Reports. 2020;8(4):461–8.

17. Rajda CM, Desabrais K, Levin MF. Relationships Between Cognitive Impairments and Motor Learning After Stroke: A Scoping Review. Neurorehabilitation and Neural Repair. 2024;39(2):142–56.

18. Winstein CJ, Stein J, Arena R, Bates B, Cherney LR, Cramer SC, et al. Guidelines for Adult Stroke Rehabilitation and Recovery: A Guideline for Healthcare Professionals from the American Heart Association/American Stroke Association. Stroke: Lippincott Williams and Wilkins; 2016. p. e98–e169.

19. Bastian AJ. Understanding sensorimotor adaptation and learning for rehabilitation. Current Opinion in Neurology. 2008;21(6).

20. Roerdink M, Beek PJ. Understanding inconsistent step-length asymmetries across hemiplegic stroke patients: Impairments and compensatory gait. Neurorehabilitation and Neural Repair. 2011;25(3):253–8.

21. Sanchez N, Schweighofer N, Finley JM. Different biomechanical variables explain within-subjects versus between-subjects variance in step length asymmetry post-stroke. IEEE Transactions on Neural Systems and Rehabilitation Engineering. 2021;29:1188–98.

22. Balasubramanian CK, Bowden MG, Neptune RR, Kautz SA. Relationship Between Step Length Asymmetry and Walking Performance in Subjects With Chronic Hemiparesis. Archives of Physical Medicine and Rehabilitation. 2007;88(1):43–9.

23. Cauraugh JH, Summers JJ. Neural plasticity and bilateral movements: A rehabilitation approach for chronic stroke. Progress in Neurobiology. 2005;75(5):309–20.

24. Hayes Cruz T, Dhaher YY. Evidence of Abnormal Lower-Limb Torque Coupling After Stroke. Stroke. 2008;39(1):139–47.

25. Sánchez N, Acosta AM, Lopez-Rosado R, Stienen AHA, Dewald JPA. Lower Extremity Motor Impairments in Ambulatory Chronic Hemiparetic Stroke: Evidence for Lower Extremity Weakness and Abnormal Muscle and Joint Torque Coupling Patterns. Neurorehabilitation and Neural Repair. 2017;31(9):814–26.

26. Sánchez N, Acosta AM, López-Rosado R, Dewald JPA. Neural constraints affect the ability to generate hip abduction torques when combined with hip extension or ankle plantarflexion in chronic hemiparetic stroke. Frontiers in Neurology. 2018;9(JUL).

27. Dewald JPA, Pope PS, Given JD, Buchanan TS, Rymer WZ. Abnormal muscle coactivation patterns during isometric torque generation at the elbow and shoulder in hemiparetic subjects. Brain. 1995;118(2):495–510.

28. Dewald JPA, Sheshadri V, Dawson ML, Beer RF. Upper-Limb Discoordination in Hemiparetic Stroke: Implications for Neurorehabilitation. Topics in Stroke Rehabilitation. 2001;8(1):1–12.

29. Perry J. Gait analysis: normal and pathological function. Second ed: CRC Press; 2010. 524–p.

30. Tyson SF, Hanley M, Chillala J, Selley A, Tallis RC. Balance Disability After Stroke. Physical Therapy. 2006;86(1):30–8.

31. Cornwell T, Finley J. Stroke impairs the proactive control of dynamic balance during predictable treadmill accelerations. Journal of The Royal Society Interface. 2025;22(231).

32. Bowen A, Wenman R, Mickelborough J, Foster J, Hill E, Tallis R. Dual-task effects of talking while walking on velocity and balance following a stroke. Age and Ageing. 2001;30(4):319–23.

33. Arene N, Hidler J. Understanding Motor Impairment in the Paretic Lower Limb After a Stroke: A Review of the Literature. Topics in Stroke Rehabilitation. 2009;16(5):346–56.

34. Van Criekinge T, Vermeulen J, Wagemans K, Schröder J, Embrechts E, Truijen S, et al. Lower limb muscle synergies during walking after stroke: a systematic review. Disability and Rehabilitation: Taylor and Francis Ltd.; 2020. p. 2836–45.

35. Moisan G, Chayasit P, Boonsinsukh R, Nester CJ, Hollands K. Postural control during quiet standing and voluntary stepping response tasks in individuals post-stroke: a case-control study. Topics in Stroke Rehabilitation. 2022;29(7):465–72.

36. Jokinen H, Melkas S, Ylikoski R, Pohjasvaara T, Kaste M, Erkinjuntti T, et al. Post-stroke cognitive impairment is common even after successful clinical recovery. European journal of neurology. 2015;22(9):1288–94.

37. Shafer RL, Solomon EM, Newell KM, Lewis MH, Bodfish JW. Visual feedback during motor performance is associated with increased complexity and adaptability of motor and neural output. Behavioural Brain Research. 2019;376:112214.

38. Agrell B, Dehlin O. The clock-drawing test. Age and Ageing. 2012;41(suppl_3):iii41–iii5.

39. Enright PL. The six-minute walk test. Respiratory care. 2003;48(8):783–5.

40. Fugl-Meyer AR, Jääskö L, Leyman I, Olsson S, Steglind S. The post-stroke hemiplegic patient. 1. a method for evaluation of physical performance. Scandinavian journal of rehabilitation medicine. 1975;7(1):13–31.

41. Berg K. Measuring balance in the elderly: preliminary development of an instrument. Physiotherapy Canada. 1989;41(6):304–11.

42. Wrisley DM, Marchetti GF, Kuharsky DK, Whitney SL. Reliability, Internal Consistency, and Validity of Data Obtained With the Functional Gait Assessment. Physical Therapy. 2004;84(10):906–18.

43. Nasreddine ZS, Phillips NA, Bédirian V, Charbonneau S, Whitehead V, Collin I, et al. The Montreal Cognitive Assessment, MoCA: A Brief Screening Tool For Mild Cognitive Impairment. Journal of the American Geriatrics Society. 2005;53(4):695–9.

44. Verghese J, Kuslansky G, Holtzer R, Katz M, Xue X, Buschke H, et al. Walking While Talking: Effect of Task Prioritization in the Elderly. Archives of Physical Medicine and Rehabilitation. 2007;88(1):50–3.

45. Van Den Bogert AJ, Geijtenbeek T, Even-Zohar O, Steenbrink F, Hardin EC. A real-time system for biomechanical analysis of human movement and muscle function. Medical and Biological Engineering and Computing. 2013;51(10):1069–77.

46. Sánchez N, Kuch A, Jeffcoat SN, Hooyman A, Haver-Hill A, Bonilla Yanez M, et al. The Differential Effects of Fast Walking Speed on Muscle Coactivation in the Paretic and Non-Paretic Extremities Post-Stroke. Neurorehabilitation and Neural Repair. 2025.

47. Rabbi MF, Pizzolato C, Lloyd DG, Carty CP, Devaprakash D, Diamond LE. Non-negative matrix factorisation is the most appropriate method for extraction of muscle synergies in walking and running. Scientific Reports. 2020;10(1).

48. Collimore AN, Aiello AJ, Pohlig RT, Awad LN. The Dynamic Motor Control Index as a Marker of Age-Related Neuromuscular Impairment. Frontiers in Aging Neuroscience. 2021;13.

49. Schwartz MH, Rozumalski A, Steele KM. Dynamic motor control is associated with treatment outcomes for children with cerebral palsy. Developmental Medicine and Child Neurology. 2016;58(11):1139–45.

50. Steele KM, Rozumalski A, Schwartz MH. Muscle synergies and complexity of neuromuscular control during gait in cerebral palsy. Developmental Medicine and Child Neurology. 2015;57(12):1176–82.

51. Friedman J, Hastie T, Tibshirani R. Regularization Paths for Generalized Linear Models via Coordinate Descent. 2010.

52. Hastie T, Tibshirani R, Friedman J. The Elements of Statistical Learning Data Mining, Inference, and Prediction. 2009.

53. Mardia KV. Measures of multivariate skewness and kurtosis with applications. Biometrika. 1970;57(3):519–30.

54. Glickman ME, Rao SR, Schultz MR. False discovery rate control is a recommended alternative to Bonferroni-type adjustments in health studies. Journal of Clinical Epidemiology: Elsevier USA; 2014. p. 850–7.

55. Lakens D. Sample Size Justification. Collabra: Psychology. 2022;8(1):33267.

56. Tibshirani R. Regression Shrinkage and Selection Via the Lasso. Journal of the Royal Statistical Society: Series B (Methodological). 1996;58(1):267–88.

57. Allen JL, Kautz SA, Neptune RR. Step length asymmetry is representative of compensatory mechanisms used in post-stroke hemiparetic walking. Gait and Posture. 2011;33(4):538–43.

58. Lewek MD, Bradley CE, Wutzke CJ, Zinder SM. The Relationship Between Spatiotemporal Gait Asymmetry and Balance in Individuals With Chronic Stroke. Journal of Applied Biomechanics. 2014;30(1):31–6.

59. Ting LH, McKay JL. Neuromechanics of muscle synergies for posture and movement. Current Opinion in Neurobiology. 2007;17(6):622–8.

60. Safavynia S, Torres-Oviedo G, Ting L. Muscle synergies: Implications for clinical evaluation and rehabilitation of movement. Topics in Spinal Cord Injury Rehabilitation. 2011;17(1):16–24.

61. Price R, Choy NL. Investigating the Relationship of the Functional Gait Assessment to Spatiotemporal Parameters of Gait and Quality of Life in Individuals With Stroke. Journal of Geriatric Physical Therapy. 2019;42(4):256–64.

62. Van Bloemendaal M, Bout W, Bus SA, Nollet F, Geurts AC, Beelen A. Validity and reproducibility of the Functional Gait Assessment in persons after stroke. Clinical Rehabilitation. 2019;33(1):94–103.

63. Kettlety SA, Finley JM, Leech KA. Visuospatial Skills Explain Differences in the Ability to Use Propulsion Biofeedback Post-stroke. Journal of Neurologic Physical Therapy. 2024;48(4):207–16.

64. McDonald MW, Black SE, Copland DA, Corbett D, Dijkhuizen RM, Farr TD, et al. Cognition in stroke rehabilitation and recovery research: Consensus-based core recommendations from the second Stroke Recovery and Rehabilitation Roundtable. International Journal of Stroke. 2019;14(8):774–82.

65. Hsu A-L, Tang P-F, Jan M-H. Analysis of impairments influencing gait velocity and asymmetry of hemiplegic patients after mild to moderate stroke. Archives of Physical Medicine and Rehabilitation. 2003;84(8):1185–93.

